# The toll of an early bell? School start time and adolescent sleep quality in Flanders, Belgium

**DOI:** 10.64898/2026.10.27.26366618

**Authors:** Ann Vandendriessche, Louise Poppe, Thijs Galmart, Margaux Rigole, Federico Bertoia, Stijn Vansteelandt, Maxim Dierckens

## Abstract

**Study objectives:** Adolescent sleep deprivation is a major public health concern, with insufficient and poor-quality sleep associated with adverse physical, mental and cognitive outcomes. Early school start times may contribute to sleep restriction by conflicting with the biologically delayed sleep patterns that emerge during puberty. This study examined the effect of school start times on sleep duration and sleep quality among secondary school students.

**Methods:** Data were obtained from the 2022 Flemish Health Behaviour in School-aged Children (HBSC) study, including 12,329 adolescents from grade 1 to 6 in 94 secondary schools in Flanders. School start times were obtained from participating schools and sleep duration and sleep quality were assessed using self-report measures. Data were analyzed using traditional statistical methods (i.e., linear regression) and debiased machine learning methods (i.e., assumption lean regression, dose response curves and modified treatment policies).

**Results:** No clinically meaningful effects of school start times on sleep outcomes were observed. In machine learning models, a 10-minute delay in start time was associated with non-significant changes in sleep quality (0.01 points on a 1-5 scale; 95% CI, −0.05 to 0.06) and school-day sleep duration (3.5 minutes; 95% CI, −1.0 to 8.0).

**Conclusions:** Within the narrow range of school start times observed in Flemish secondary schools, confidence intervals excluded clinically meaningful effects of later start times on adolescent sleep duration and sleep quality. Whether larger delays would improve sleep remains to be determined.

**Statement of significance:** Adolescent sleep deprivation is a major public health concern, and delaying school start times has been widely advocated as a structural strategy to improve sleep. However, evidence remains largely observational, with varying approaches to confounding control, and European data are scarce. Using causal inference methods in a population-based sample of over 12,000 Flemish secondary school students, this study showed that differences in school start times within the range currently observed were unlikely to meaningfully improve sleep duration or sleep quality. These findings suggest that modest variations in start times may be insufficient to improve adolescent sleep, highlighting the need to evaluate whether substantially later start times, while accounting for factors like commuting time, are required to generate measurable sleep benefits.

## Introduction

Chronic sleep deprivation among adolescents is a major public health concern, both internationally [1] and in Flanders [2, 3]. Recent Flemish data indicate that 67% of adolescents aged 13–18 years fail to meet the National Sleep Foundation’s [4] recommended minimum of 8 hours of sleep per night, and nearly half report poor sleep quality, difficulties falling asleep and morning fatigue [2]. Adequate sleep is essential for adolescents to support optimal physical health, emotional regulation and cognitive functioning. Insufficient and poor-quality sleep during adolescence has been associated with increased risks of stress, anxiety and depressive symptoms [5–7], as well as obesity, type 2 diabetes and hypertension [8, 9]. In addition, inadequate sleep impairs attention, executive functioning and memory, negatively affecting school performance [10].

A central mechanism underlying adolescent sleep restriction is the biological–social mismatch that emerges during puberty [11]. Biological changes in circadian timing shift sleep patterns toward a later phase, leading adolescents to naturally feel alert later at night and to prefer later wake times [11, 12]. Societal demands such as early school start times remain however unchanged, creating a misalignment between biological and social timing [11]. This misalignment contributes to shortened sleep duration on school days and the accumulation of sleep debt. Many adolescents compensate by sleeping longer during weekends, further shifting their sleep timing and making it harder to fall asleep early before the start of a new school week [13]. This recurring shift in sleep timing between school days and non-school days is commonly referred to as “social jetlag” [14]. Moving beyond individual behaviors that negatively influence sleep such as screen use and caffeine consumption [15], school start times represent a structural factor that might systematically constrain adolescents’ opportunity to obtain sufficient and restorative sleep.

Delaying school start times has been proposed as a potential strategy to mitigate chronic sleep restriction [16]. Reflecting growing concern about insufficient adolescent sleep, the American Academy of Pediatrics recommended that secondary schools start no earlier than 8:30 a.m [17]. A meta-analysis indeed indicated that later start times are generally associated with longer sleep duration, reduced daytime sleepiness and improvements in mood and alertness [18]. However, most of the available evidence originates from the United States, where school systems and daily schedules differ from those in Europe (i.e. earlier start times, longer commuting distances to school, more passive transport, differently organized extracurricular activities) [19–22]. In addition, because cluster-randomized controlled trials of school start times are difficult to implement, the existing evidence base relies largely on observational studies, many of which have been limited by inadequate identification and control of confounding factors [20, 23]. Strengthening causal inference from observational data requires careful identification and control of confounding factors based on substantive knowledge and explicit causal assumptions [24]. Together, these contextual and methodological limitations restrict the generalizability and interpretability of existing findings, highlighting the need for more rigorous evidence in European settings. In Flanders, secondary schools typically start around 8:20–8:30 a.m., but the potential causal effects of variation in school start times on adolescent sleep have not yet been thoroughly examined.

The present study aims to address these gaps by providing rigorous, data-driven evidence on the causal effect of school start times on sleep duration and sleep quality among adolescents, using data from the 2022 Flemish wave of the Health Behaviour in School-aged Children (HBSC) study. Compared with much of the existing literature, this population-based dataset provides relatively rich information on individual-, family- and school-level characteristics that may be relevant to confounding. Informed by a prespecified directed acyclic graph (DAG), this information enabled a systematic approach to confounder selection. This theory-driven causal framework was combined with both conventional statistical models and modern debiased machine learning methods.

## Methods

### Study design and participants

This cross-sectional study used data from the 2022 Flemish wave of the HBSC study, an international survey conducted every four years in collaboration with the World Health Organization Regional Office for Europe. The HBSC study adheres to a standardized international research protocol and collects nationally representative data on health behaviors, lifestyle factors and well-being among adolescents [25]. In Flanders, HBSC data are collected in both primary and secondary education. Standard HBSC age groups include students aged approximately 11, 13 and 15 years, supplemented in Flanders by additional grades to cover the broader age range of 10 to 18 years. Because the present study focuses on the impact of school start times in the context of pubertal changes in sleep–wake regulation, only students attending secondary education were included in the analyses.

Data collection took place between February and June 2022 using a two-stage cluster sampling design. First, schools were randomly selected from the official registry of Flemish schools, stratified by educational network to ensure representativeness. Second, classes within participating schools were randomly selected based on class lists containing information on grade, sex distribution and educational track. Of the 697 invited secondary schools, 103 participated, yielding data from 16,193 students drawn from an initial sample of 23,759 selected students. Questionnaires were administered during regular class hours under supervision of teachers or school staff, either online or in paper-and-pencil format. Participation was voluntary, questionnaires were processed anonymously and passive parental consent was obtained [25, 26]. The study was in accordance with the Declaration of Helsinki and ethical approval was granted by the Ethics Committee of the University Hospital Ghent (2013/1145 BC-00425).

Information on school start times during the 2022 academic year was collected retrospectively between December 2024 and February 2025. All secondary schools (n=103) were contacted by e-mail and invited to complete a short online questionnaire on school start times. Non-responding schools were contacted by telephone to obtain the same information. Students with missing data on relevant variables (n = 2,505) or invalid class identifiers (n = 16) were removed. Students attending schools with varying start times across days were excluded (n = 1,210) because a unique school start time could not be assigned. Finally, students reporting implausible sleep durations (less than 3 or more than 14 hours) were excluded (n = 133). The final analytical sample comprised 12,329 students nested within 94 schools (nine schools were excluded during preprocessing due to missing data or varying start times across days).

### Measures

#### School start times

were asked via an online questionnaire among all participating secondary schools. Schools were first asked whether the official start time (i.e., the time students were expected to be present at school) was identical for all students across grade levels and days of the week. If so, schools were asked to report the exact daily start time in hh:mm format. If start times varied by grade level or by day (e.g., alternating schedules), schools were asked to describe these arrangements in an open-response field.

#### Sleep duration

on school and weekend days was calculated using four self-reported items: usual bedtime, usual time attempting to fall asleep, sleep latency and usual wake-up time [27], a method recommended in the sleep literature and shown to improve the validity of self-reported sleep duration compared with objective measures [28] and more accurately estimating self-reported sleep [3]. Bedtime and wake-up time were reported in 30-minute intervals; sleep latency was reported in 10-minute categories. Sleep duration was computed by subtracting sleep latency from the interval between reported bedtime and wake-up time.

#### Sleep quality

was assessed using five items derived from the Groningen Sleep Quality Scale [29], an instrument validated among adults with good internal consistency. Items assessed perceived poor sleep, difficulty falling asleep (>30 minutes), insufficient sleep, nocturnal awakenings and feeling rested upon waking. Responses were rated on a five-point Likert scale ranging from “never” (1) to “always” (5). The item assessing feeling rested was reverse coded, and items were summed and divided by five to create a total sleep quality score (ranging from 1 to 5), with higher scores indicating poorer sleep quality.

#### Covariates

Potential confounders were identified a priori using a Directed Acyclic Graph (DAG) [30] based on existing literature, expert knowledge and the available data [31] (Figure 1). Variables that were considered relevant but that were not measured (commuting time, support staff-to-student ratio, teacher-to-student ratio, and school regulations on digital communication via school platforms) are shown in the DAG as unmeasured nodes. The DAG assumes that school start time influences school choice. Because the reverse direction is also conceivable, the DAG should be interpreted within this framing. Individual-level covariates included age, sex, and socioeconomic status as measured by family affluence level, parental education level, and parental employment. School-level covariates included the type of education (A or B-type, general, technical, arts or vocational), educational network (privately run, community, and officially funded), school-level educational poverty indicator, and urbanicity (inhabitants per square kilometer).

**Figure 1.**
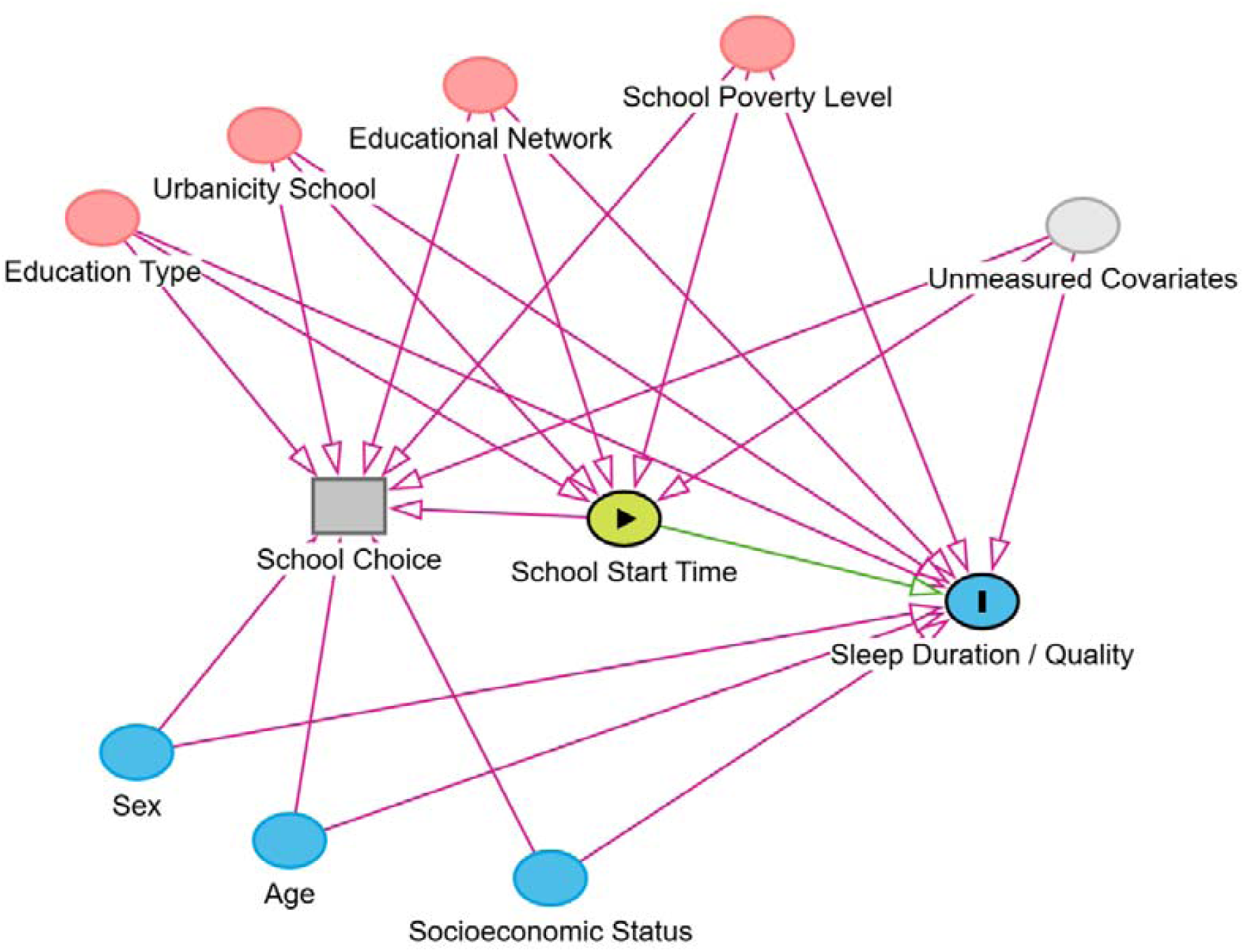
Directed Acyclic Graph (DAG) for the causal effect of school start times on adolescent sleep outcomes. ***Note***. To improve readability, causal relationships between confounding variables are omitted and should not be interpreted as assumptions of independence. Parental education, parental employment, and family affluence level were grouped into *Socioeconomic Status* to improve readability of the DAG. Variables that were not measured (commuting time, support staff-to-student ratio, teacher-to-student ratio, regulations on digital communication) are shown as *Unmeasured Covariates*.

### Statistical analysis

Four complementary methods were used to evaluate the association between school start times and the sleep outcomes. This approach adds value by progressively relaxing parametric assumptions to reduce potential model misspecification bias. It furthermore adds value by evaluating the effect of realistic, individualized shifts (or modified treatment policies, MTPs) in school start time, as opposed to contrasting uniform start times. Table 1 provides an overview of the estimands targeted by each considered method.

**Table 1.** Overview of the statistical models and their estimands.

| Model | Method | Estimand |
| --- | --- | --- |
| 1 | Ordinary Least Squares regression | The effect on the expected outcome of shifting the start time by one unit, assuming a correctly specified linear model |
| 2 | Assumption-lean estimation | The (best linear approximation of the) effect on the expected outcome of shifting the start time by one unit |
| 3 | Continuous dose-response curve estimation | The expected mean outcome if everyone received the exact same start time |
| 4 (a-d) | Modified Treatment Policies | The effect on the expected outcome of shifting the start time according to a pre-specified policy |

First, Ordinary Least Squares (OLS) regression [25, 26] was used to estimate the linear association between school start time and sleep outcomes. OLS assumes a correctly specified strictly linear model and was implemented to include all measured covariates as main effects. As such, it serves as a parametric benchmark against which the results from more flexible methods can be compared. Second, to reduce potential model misspecification from the OLS model, an assumption-lean estimator [32, 33] was used. This method uses an ensemble of parametric and machine learning approaches, also known as Super Learner [34], to flexibly model the relation between confounders and outcome, and between confounders and school start time. By avoiding strict parametric assumptions, this method provides a robust summary measure of the linear association. Third, this linearity is further relaxed using kernel smoothing, resulting in continuous dose-response curves estimated using the *npcausal* package in R [35]. Finally, the effects of four realistic MTPs on the outcomes were evaluated using the *lmtp* package in R [36–38], again using an ensemble of parametric and machine learning approaches for flexible modeling of the relation between confounders on the one hand, and outcome and school start time on the other hand. Unlike static interventions that assign all individuals to the same exposure level regardless of feasibility, MTPs define interventions relative to each individual’s observed exposure level. Specifically, four interventions based on observed school start times were evaluated: (a) advancing all start times before 8:30 a.m. to 8:30 a.m.; (b) delaying start times by 5 minutes; (c) delaying start times by 10 minutes and (d) delaying start times by 30 minutes. For the latter three MTPs, delayed start times were capped at 9:00 a.m. to preserve the feasibility and policy relevance of the hypothetical intervention. Given that Flemish secondary schools typically start around 8:20 – 8:30 a.m., a 9:00 a.m. start time represents a substantial but still plausible delay within the educational context studied.

Across all models, all measured confounders were adjusted for, and standard errors were calculated, accounting for clustering at the class level to account for the nested data structure (except for the continuous dose-response curves). Clustering at the classroom level was chosen because sleep behaviors may be more strongly correlated among students within the same class. The sampling design was stratified by educational network. Educational network was included as a covariate in the linear regression (Model 1) and the assumption-lean estimator (Model 2), which accounts for the stratification in these models. In the dose-response curves (Model 3) and the MTPs (Model 4), educational network was entered only as one of the confounders in the flexible nuisance models, and the stratified design was not otherwise taken into account. All statistical analyses were performed using R [39].

## Results

### Sample characteristics

The study sample (n = 12,329) consisted of an equal portion of boys (50.4%) and girls (49.6%) with a mean age of 15.48 years (standard deviation: 3.3, range: 9.5 – 22.5). The average school start time was 8:26 a.m. (range: 8:00 a.m. – 9:00 a.m.; Figure 2). The mean sleep quality was 2.91 on a five-point scale (standard deviation: 0.74, range: 1 – 5), with higher scores indicating poorer sleep quality. The mean sleep duration on school days was 7.4 hours (standard deviation: 1.5, range: 3.0 – 11.9), and 9.1 hours on weekend days (standard deviation: 2.3, range: 3.0 – 13.9).

**Figure 2.**
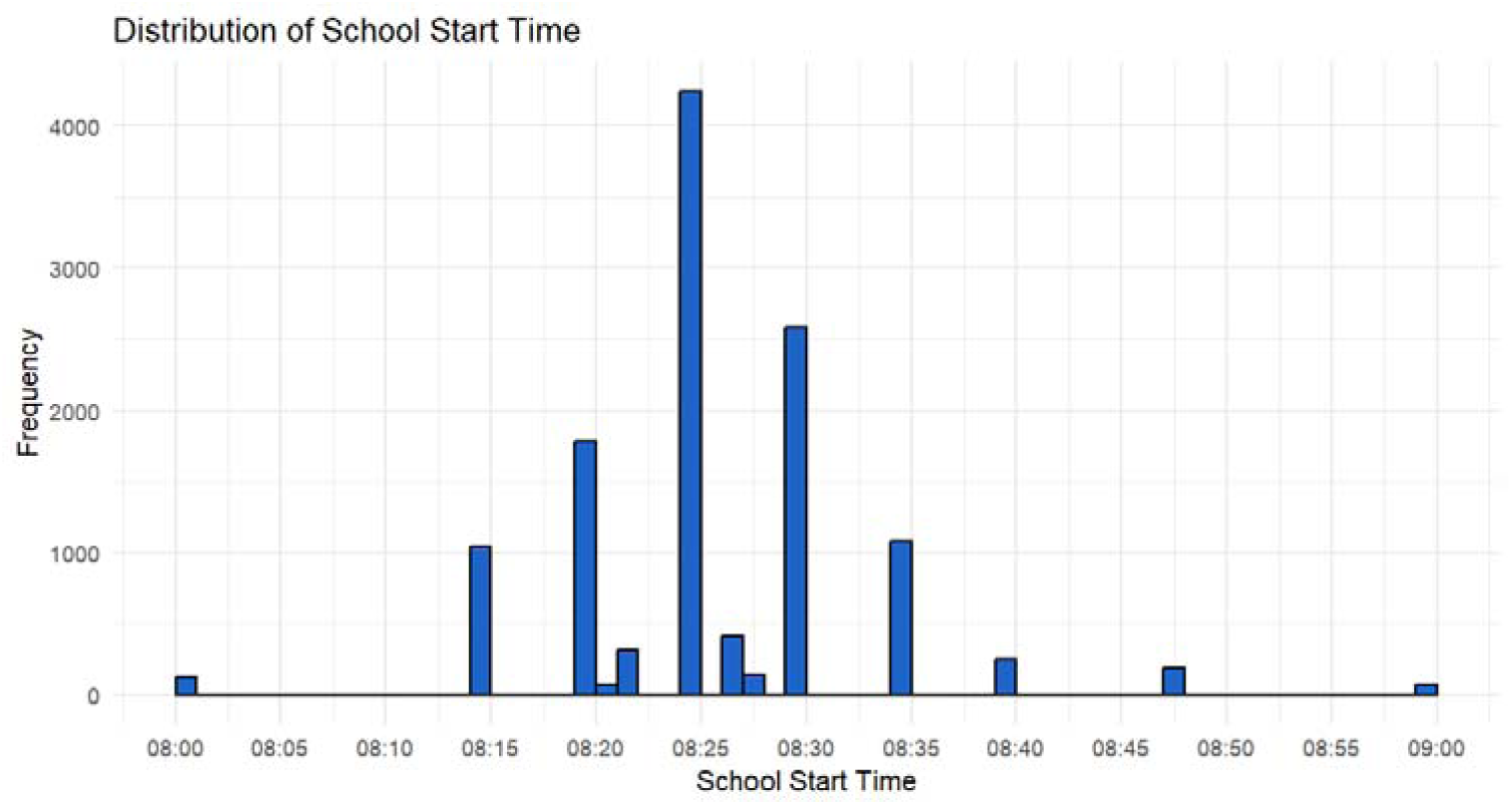
Distribution of the school start times at the student level (n = 12,329).

Mean school urbanicity was 1,257 inhabitants per square kilometer (standard deviation: 5,210 range: 112 – 18,633), the mean school poverty level was 0.89 on a 0 to 4 scale with higher scores indicating greater poverty (standard deviation: 0.26, range: 0.19 – 3.11), and the mean Family Affluence Scale score was 9.4 on a 0 to 13 scale with lower scores indicating lower affluence (standard deviation: 3.8, range: 0 – 13). Education types were distributed across general (35.0%), A-type (29.4%), technical (18.7%), vocational (13.0%), B-type (2.2%), and arts (1.7%) secondary education. Educational networks comprised privately run (69.0%), community (25.9%), and officially funded (5.1%) institutions. For highest parental education, 62.4% had completed university, 15.0% secondary school, 1.1% basic education, 0.7% had never studied, and 20.8% were unknown by the students. Parental employment status was 97.6% employed, 2.0% unemployed, and 0.4% unknown by the students.

### School start time and sleep quality

No evidence for a meaningful effect of a later school start time on sleep quality was found. In the standard linear regression model (Model 1), a 10-minute delay in start time was associated with an estimated change of −0.01 points (on a scale from 1 to 5) in sleep quality (95% CI, −0.03 to 0.02). This finding persisted when applying the debiased machine learning estimator (Model 2) to account for complex confounding, which yielded an estimate of 0.01 points (95% CI, −0.05 to 0.06). The dose-response curve (Model 3; Figure 3, panels A1 and A2) indicated little variation in sleep quality across the full range of school start times. Results from the MTP analyses (Models 4a – 4d) were consistent with these findings. Standardizing all school start times earlier than 8:30 a.m. to 8:30 a.m. yielded an estimated sleep quality change of 0.00 points (95% CI, −0.01 to 0.02). Incremental delays of 5 and 10 minutes were associated with estimated changes of −0.01 (95% CI, −0.02 to 0.01) and −0.01 points (95% CI, −0.03 to 0.01), respectively. A 30-minute delay (capped at 9.00 am), resulted in an estimated change of −0.04 (95% CI, −0.06 to −0.02). Although this change is significantly different from zero (improved sleep quality), the boundaries of the confidence interval suggest very small potential changes relative to the five-point sleep quality scale, providing little practical and clinical relevance.

**Figure 3.**
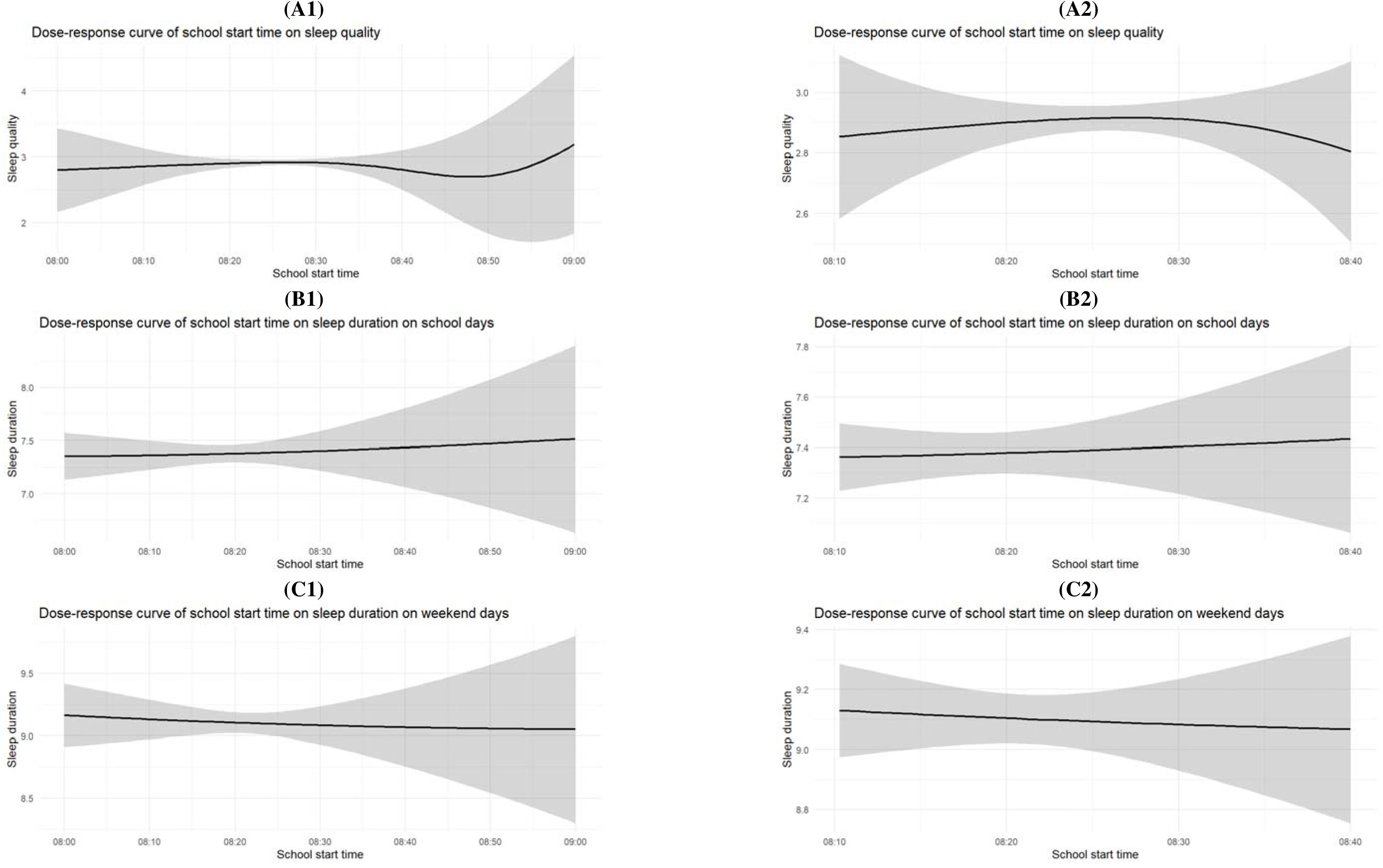
Continuous dose-response associations between school start times and adolescent sleep outcomes. ***Note.*** Panels present the estimated associations for sleep quality (A1, A2), sleep duration on school days (B1, B2), and sleep duration on weekend days (C1, C2). Shaded areas represent 95% confidence intervals. Panels labelled ‘2’ provide enlarged views of the corresponding panels labelled ‘1’.

### School start time and sleep duration on school days

No evidence for a meaningful effect of a later school start time on sleep duration on school days was found. The linear model (Model 1) indicated a marginal increase of 1.3 minutes of sleep per 10-minute delay in school start time (95% CI, −0.7 to 3.4), while the debiased machine learning approach (Model 2) estimated an increase of 3.5 minutes (95% CI, −1.0 to 8.0). The dose-response analyses (Model 3; Figure 3, panels B1 and B2) similarly indicated little variation in sleep duration across the observed range of school start times. MTP analyses (Models 4a – 4d) similarly showed negligible gains in sleep duration. Standardizing all start times earlier than 8:30 a.m. to 8:30 a.m. resulted in an estimated change of 0.0 minutes (95% CI, −1.6 to 1.6). Delays of 5 and 10 minutes yielded non-significant increases of 0.2 minutes (95% CI, −1.3 to 1.7) and 0.3 minutes (95% CI, −1.4 to 2.0), respectively. A 30-minute delay (capped at 9:00 a.m.) was associated with an estimated increase of 1.0 minute (95% CI, −0.8 to 2.7).

### School start times and sleep duration on weekend days

No evidence for a meaningful effect of school start time on sleep duration during weekend days was found. The linear (Model 1) and debiased machine learning (Model 2) models estimated changes of 1.3 minutes (95% CI, −1.2 to 3.7) and 3.5 minutes (95% CI, −2.3 to 9.4) additional sleep per 10-minute delay in school start time, respectively. The dose-response analyses (Model 3; Figure 3, panels C1 and C2) again indicated little variation in sleep duration across the observed range of school start times. MTPs projected similarly small changes: standardizing all start times earlier than 8:30 a.m. to 8:30 a.m. yielded an estimated difference of 1.1 minutes (95% CI, −0.9 to 3.0). Delays of 1 and 10 minutes were associated with estimated changes of 0.5 minutes (95% CI, −1.3 to 2.3) and 0.9 minutes (95% CI, −1.1 to 2.9), respectively. A 30-minute delay (capped at 9:00 a.m.) resulted in a weekend sleep increase of 1.9 minutes (95% CI, −0.2 to 4.1).

## Discussion

The present study examined the causal effect of school start times on sleep duration and sleep quality among adolescents using data from the 2022 Flemish HBSC study. Overall, the findings provide no evidence that variation in school start times substantially affects adolescent sleep outcomes. Later school start times were associated with only marginal improvements in sleep duration and sleep quality, and these effects were unlikely to be of practical or clinical relevance.

These findings differ from much of the international literature, which generally reports beneficial effects of later school start times on adolescent sleep and daytime functioning [18–21, 40]. However, reported effect sizes vary considerably across studies and settings. For example, the Canadian HBSC study found that a 10-minute delay corresponded to approximately 3.2 [95% CI, 2.0 to 4.5] additional minutes of sleep per night [31], while positive effects have also been observed following delays of 15 [41] and 30 minutes [42]. At the same time, some studies suggest that delays of up to 20 minutes may be insufficient to produce meaningful improvements [43]. The limited effects observed in the present study may therefore reflect the relatively narrow range of school start times observed in Flemish secondary schools rather than a contradiction of previous research. Most schools in the present study started between 8:20 and 8:30 a.m., and only a small number (n=12) began after 8:30 a.m. (seven of them at 8:35 a.m.), the threshold recommended by the American Academy of Pediatrics [17]. Consequently, the study primarily captures relatively small differences in school start times, and estimates for substantially later start times are based on comparatively little information. This may have constrained the ability to detect larger effects and may partly explain why the observed associations were smaller than those reported in studies examining more substantial changes in school start times. The findings therefore suggest that the modest differences in school start times are unlikely to meaningfully affect adolescent sleep in the Flemish context, but should not be interpreted as evidence that larger delays would be ineffective.

The potential effect of a given school start time may depend not only on the clock time itself, but also on contextual factors such as commuting patterns, local school schedules [19–22], daylight conditions and the timing of sunrise [40]. Recent evidence suggests that identical nominal school start time may have different biological implications across geographical and environmental contexts [40]. Such contextual differences may therefore contribute to the heterogeneity in findings between studies and countries.

Differences between the present findings and previous studies may also reflect variation in study design and confounding control. Adjustment strategies varied considerably across previous studies. Some studies relied on uncontrolled pre-post evaluations [42, 44], whereas others adjusted for a limited set of individual characteristics such as age, sex, family income or mother tongue [41, 43]. Gariepy et al. [31] used a more extensive adjustment set, including grade, family affluence and several school-level geographic characteristics (e.g., rurality, latitude and province), whereas Widome et al. [45] adjusted for multiple individual- and family-level characteristics and accounted for clustering within schools. In contrast, the present study used a prespecified DAG to identify potential confounders and incorporated a broader range of individual-, family- and school-level characteristics, including age, sex, family affluence, parental education and employment, as well as type of education, educational network, school-level educational poverty and urbanicity. Although residual confounding by unmeasured variables cannot be ruled out (cfr. *Infra*), this approach strengthens the causal interpretation of the findings.

Measurement characteristics may also have attenuated detectable effects. Sleep duration was derived from self-reported bedtimes, sleep onset times and wake-up times, collected in 30-minute response intervals. These broad categories may obscure subtle differences in sleep duration associated with small variation in school start times. In addition, behavioral adaptation could further contribute to reduced observable effects. Adolescents may compensate for later wake-up times by going to bed later. Although delayed start times have been associated with slightly later bedtimes [44], previous research indicates that these shifts are generally smaller than the corresponding changes in wake-up time, resulting in a net increase in sleep duration [40, 44, 45]. Bedtime compensation is therefore unlikely to fully offset the potential benefits of later school start times, although its role in the Flemish context remains uncertain.

The present study has several strengths. To our knowledge, it is the first study to examine school start times and adolescent sleep in Flanders using a causal inference approach, thereby strengthening the interpretation of our findings by explicitly addressing potential confounding and providing a more robust estimate of the association between school start times and sleep duration than would be possible using conventional observational analyses. Furthermore, the analyses were based on a large, representative population-based HBSC sample, providing robust coverage of Flemish secondary school students. Several limitations should nevertheless be acknowledged. First, all sleep measures were based on self-report and may be affected by measurement error, including recall bias and reporting inaccuracies [46]. Second, the causal interpretation of the estimates relies on the assumption that there is no unmeasured confounding, which cannot be verified from the data. The DAG (Figure 1) included several variables that would ideally have been available but were not measured, including commuting time, support staff-to-student ratio, teacher-to-student ratio, and school regulations on digital communication outside school hours. Commuting time may be particularly relevant as it could influence both school choice and sleep behavior, the latter of which has been documented [47]. Although no direct evidence was identified linking commuting time with school choice or school start times, it is plausible that commuting time is related to school choice and may therefore act as an unmeasured confounder. While differences between rural and urban schools (captured by the urbanicity covariate) may partially reflect variation in commuting patterns, individual commuting time was not available and may still have influenced the observed associations. If longer commuting times were more common among students attending later-starting schools, and longer commuting times reduced sleep duration, failure to account for commuting time could have attenuated the observed association between school start times and sleep. Third, schools were sampled stratified by educational network. Although this was addressed through covariate adjustment in Models 1 and 2, it was not explicitly accounted for in Models 3 and 4, and its potential impact on the estimates was not evaluated. The standard errors and confidence intervals of Models 3 and 4 may therefore not fully reflect the sampling design.

Future research should build on the present findings and evaluate more substantial delays in school start times than are currently observed in Flemish secondary schools, preferably using observational or quasi-experimental studies and more precise sleep measures. Particular attention should be paid to contextual factors including commuting patterns, local daylight conditions and the alignment between school start times and solar time, as these factors may shape the effectiveness of later school start times and thereby inform decisions regarding optimal school scheduling. Future research should therefore investigate how these contextual factors influence the effects of school start times across different settings.

## Conclusion

This study found no evidence that variation in school start times within the range currently observed among Flemish secondary schools has a meaningful effect on adolescent sleep duration or sleep quality. Although estimated effects generally pointed towards slightly improved sleep outcomes with later school start times, their magnitude was small and unlikely to be practically or clinically relevant. While these findings differ from much of the international literature, they may partly reflect the limited variability in school start times examined in the present study. Differences in study design and the extent to which potential confounders were identified and controlled may also contribute to variation in findings across studies. Further research should evaluate whether more substantial delays in school start times would yield measurable benefits for adolescent sleep in Flanders and further strengthen the causal evidence base on the effects of school start times on adolescent sleep outcomes.

## Data Availability

The datasets used and/or analysed during the current study are available from the corresponding author on reasonable request.

## Acknowledgements

The authors would like to thank the participating schools and students of this study.

## Author contributions

A.V. (Conceptualization, Methodology, Writing – Original Draft, Writing – Review & Editing, Supervision, Funding Acquisition), L.P. (Conceptualization, Methodology, Data Curation, Writing – Review & Editing), T.G. (Methodology, Formal Analysis, Data Curation, Writing – Original Draft, Writing – Review & Editing, Visualization), M.R. (Data Curation, Writing – Review & Editing), F.B. (Methodology, Writing – Review & Editing), S.V. (Methodology, Writing – Review & Editing), and M.D. (Conceptualization, Methodology, Data Curation, Writing – Review & Editing, Supervision, Project Administration, Funding Acquisition).

